# ALBUMIN CREATININE RATIO IN TYPE 2 DIABETES MELLITUS IN BARBADOS: ALBUMIN CREATININE RATIO AS AN INDICATOR OF NON-HEALING FOOT WOUND FORMATION IN DIABETES

**DOI:** 10.1101/2021.10.01.21264433

**Authors:** André R Greenidge, Kim R Quimby, Amy P Speede, Ian R Hambleton, Simon G Anderson, R Clive Landis

## Abstract

**Aims:** To investigate whether kidney injury, determined by albumin creatinine ratio, was associated with current non-healing foot wounds in type 2 diabetes.

**Materials and Methods:** Eighty-nine Barbadians with diabetes were recruited. Cases had a current foot wound and controls had no current foot wound and no history of a non-healing foot wound. Cases were matched to controls using sex, age and duration of diabetes. Participants were from wound dressing and diabetes clinics at the Queen Elizabeth Hospital and Polyclinics, and from private healthcare practitioners. The relationship between albumin creatinine ratio and foot ulceration, adjusting for selected potential risk factors, was analyzed using logistic regression and presented as odds ratios.

**Results:** Forty-four cases and 45 controls were matched, with no statistically significant difference in matching criteria. There were statistically important differences in measures of neuropathy, blood glucose, HbA1c and Albumin:creatinine ratio between cases and controls. Cases were 3 times more likely than controls to have microalbuminuria (95% CI 0.9 – 10.2; p=0.08). Cases were 7.4 times more likely than controls to have macroalbuminuria (95% CI 1.2 – 47.5; p=0.04).

**Conclusions:** The possible association of albumin:creatinine ratio with diabetic foot wounds raises the possibility of its use in earlier identification of persons on the pathway to developing diabetic foot.

## INTRODUCTION

Diabetes mellitus is affecting a rapidly rising proportion of the globe’s population from an estimated 171 million persons living with diabetes worldwide in 2002 to 463 million in 2019^1;2^ (1; 2).

The high levels of circulating plasma glucose in diabetes affects tissue and organ systems within the body, often leading to many complications such as diabetic foot ulceration. In Barbados, many of these ulcers present as chronic non-healing foot ulcers which ultimately lead to amputation^3^ (3). Diabetic foot syndrome, including lower-extremity amputations is a major contributor to the mortality and morbidity from diabetes in developed countries^4^ (4). In the developing world, evidence suggests that all-cause mortality after diabetes related lower extremity amputation is considerably higher than in developed countries^5:6^ (5; 6). The higher mortality rate was demonstrated in the Wellcome Diabetic Foot study in Barbados which revealed the second highest prevalence of diabetic lower extremity amputations in females reported globally, and where the full consequence of amputation in the population was borne out with 66% mortality in the first year subsequent to an above the knee amputation^3;7^ (3; 7).

With such clear evidence of the consequence of amputation in our population, investigations into diabetic foot are warranted and necessary. However, much of the global work on diabetic foot ulceration focuses on amputations, by which time it is too late to intervene and change the course of disease progression. For more timely assessment and interventions, early markers of disease presence and progression should be investigated. One early marker of diabetic foot that must be considered is diabetic neuropathy. Neuropathy is currently the best independent marker of diabetic foot, with studies indicating that having a vibration perception of above 25 volts shows an increased risk of developing diabetic foot ulcers^8^ (8). Neuropathy is characterized by damage and degeneration of the peripheral nerves and the resultant loss of nerve function and pain sensation leads to increased risk of injury and trauma to the feet^8-11^ (8-11). Neuropathy has been shown to be a risk factor for developing diabetic foot ulceration and around 15% of persons with diabetes will suffer with some form of ulceration in their lifetime^8;12^ (8; 12). However, much like amputations, clinically significant neuropathy may present in more advanced stages of diabetes progression, when effective intervention is not possible^13^ (13).

Another factor in the equation of diabetic foot wounds, is deranged wound healing. In normal, uncompromised wound healing, the ultimate aim of the process is to repair the damage and return the tissue to its fully functioning previous state. The process can be broken down into two phases; a pro-inflammatory phase and an anti-inflammatory / wound healing phase. The pro-inflammatory phase occurs up to 72 hours after tissue injury, includes inflammation and is directed by the presence and release of pro-inflammatory cytokines and growth factors^14^ (14). After the pro-inflammatory phase, there is an orderly progression through the inflammatory phase, and ultimately a transition to the anti-inflammatory / wound healing phase. It is in this phase that tissue regeneration and repair occurs. The director and key cell in this process is the macrophage. Macrophages have two main phenotypes and they exhibit a plasticity in how they can switch between phenotypes depending on the circumstance. In this plasticity, macrophages can switch from the M1 inflammatory phenotype, where killing and destruction is promoted, to the M2 anti-inflammatory phenotype, where growth and healing is promoted^15^ (15). Studies have indicated that persistent inflammation, directed by macrophages, is at the root of impaired wound healing in both animal and human models of diabetes^16-19^ (16-19).

Previous studies have explored an association between kidney dysfunction and diabetic foot. Some investigations have revealed that undergoing dialysis was associated with foot ulceration in persons with stage 4 or 5 chronic kidney disease while other studies have indicated that albuminuria is more prevalent in persons with diabetic foot and is also a predictive factor of in-hospital mortality^20-23^ (20-23). Mechanistically, we have proposed that the cellular basis of nephropathy may be linked with non-healing wounds in diabetic foot through a shared pathogenesis in which macrophages perpetuate runaway inflammation and tissue remodeling^24^ (24). In this Wound Healing Study (WHY) in diabetes, we examined neuropathy and renal injury markers in a case-control study of persons with type 2 diabetes, to investigate any association with early stage kidney disease and early phase impaired wound healing in diabetes.

## MATERIALS AND METHODS

### Study Approval

All human investigations in this study were conducted according to the principles expressed in the Declaration of Helsinki. Ethical approval was granted from both the Ministry of Health and the Institutional Review Board of the Ministry of Health and the University of the West Indies. All recruited participants were free to terminate their participation in the study at any time with the full understanding that future medical care would not be affected.

### Study Design

Determining kidney injury by albumin: creatinine ratio was one component of the WHY study. The WHY study was a case-control study, with both cases and controls having diabetes. Cases were persons of African descent, living with type 2 diabetes, with a current non-healing (30+ days) foot ulcer below the malleolus of the foot. Controls were persons of African descent, living with type 2 diabetes but having no current and no previous history of a non-healing (30+ days) foot ulcer below the malleolus of the foot. The study design created 1:1 case-control matching by age, sex, and diabetes duration. Primary endpoints of the WHY study were of a genetic nature, examining the proportions of different haptoglobin phenotypes and TNF Receptor Associated Periodic Syndrome mutations in the case and control populations. Exclusion criteria for the study included if the participant; was not resident in Barbados for >1 year, had no medical professional diagnosis of type 2 diabetes, did not self-identify as being of African descent, had no current ulcer but did have a history of non-healing foot ulcer, currently had cancer, currently had connective tissue diseases such as lupus or rheumatoid arthritis or scleroderma, currently had a blood disorder such as sickle cell disease. Participants were also excluded if they had a current metabolic condition such as gout or amyloidosis, if they had a currently inflammatory skin condition such as panniculitus, pyoderma gangrenosum or psoriasis. An immunodeficiency condition, medical professional diagnosed kidney disease and or renal replacement therapy were also reasons for exclusion from the study.

### Participant Recruitment

The study population was recruited with informed consent from the wound dressing and diabetes clinics of the public Queen Elizabeth Hospital, Barbados’ main public hospital, from Polyclinics, which are secondary healthcare centers on the island, as well as the clinics of private healthcare practitioners. Recruitment occurred between July 2010 and December 2013. Forty-four (44) cases and forty-five (45) controls were matched for age (to within 5 years), sex and duration of diabetes (<5 years, 5-10 years, 10+ years).

### Participant Measurement

Anthropometric measurements were taken from all participants. The standing weight of the Participant was taken with the participant standing on a SECA 750 scale (Seca Corporation. Chino, CA. USA) with the measurement taken to the nearest 0.1kg. The standing height of the participant was taken with the participant standing on a SECA 213 stadiometer (Seca Corporation) with the measurement taken to the nearest 0.5cm. The standing hip circumference of the Participant was taken with the participant standing on the floor and with the aid of a tape measure to the nearest 0.5cm. This was similar to the method of measuring the participant’s waist circumference. Blood pressure was measured at three intervals after 5-10 min of rest in a supine position using a standard mercury sphygmometer (W.A. Baum Co. Inc, NY, USA). Vibration perception threshold (VPT) measurements were obtained using a neurothesiometer (Horwell Scientific Laboratory Supplies, Nottingham, UK). Two modes of stimulation were used; first the intensity of the stimulus on the plantar surface of the foot was increased until it was perceived and second the intensity of the stimulus was decreased until it was not perceived. The lesser of the two values was used if they differed.

### Biological Sample Testing

Fasting EDTA blood samples were obtained from participants. Fasting blood clinical test levels were assessed by using the Roche Reflotron Plus Analyzer (Roche Diagnostics GmbH, Mannheim Germany) according to the manufacturer’s instruction for the measurement of glucose, triglycerides, HDL cholesterol, total cholesterol and the calculation of LDL cholesterol. Urinary albumin/creatinine ratio as well as HbA1c were assessed using the respective kits of the Bayer DCA 2000+ Analyzer (Bayer HealthCare LLC Elkhart, IN. USA) according to the manufacturer’s instruction.

### Statistical Analysis

All analyses were facilitated by the paired design of the study. All tests were two-sided, and the nominal level of α was 5%. The consideration used was that albumin: creatinine ratio in cases would be significantly higher than controls. The sample size was calculated as a difference between two means, and sample size calculations indicated that a minimum of 35 cases and 35 controls would allow a paired t-test to have 80% power to distinguish between the groups when they differed by at least 10%. The analysis of the data generated Odds Ratios (OR), both univariate and then adjusted for age, sex and duration of diabetes. Conclusions were based on adjusted analyses. All results were compiled and analysis performed using STATA 14 (StataCorp LP, College Station, TX. USA).

## RESULTS

The demographic data collected from the study participants is presented in **Table 1**. All subsequent variables in the analysis are presented as odds ratios between cases and controls, both univariate and adjusted for age, sex and duration of diabetes where appropriate.

**Table 1.** Demographic Data of WHY Study Participants.

| **VARIABLE** | **CASE [44]** | **CONTROL [45]** | **UNIV. OR (95% CI)** | **ADJ 1<sup>†</sup> OR (95% CI)** | **P** |
| --- | --- | --- | --- | --- | --- |
| *Age (years)* | 58.4 | 58.3 | 1.00 (0.96 – 1.04) | 0.99 (0.95 – 1.04) | 0.79 |
| *Sex (male) %* | 40.0 [18] | 40.9 [18] | 1.04 (0.45 – 2.42) | 0.89 (0.38 – 2.11) | 0.79 |
| *DM Duration (years)* | 15.2 [43] | 12.8 [45] | 1.02 (0.98 – 1.06) | 1.02 (0.98 – 1.06) | 0.31 |
| *Age DM onset (years)* | 43.3 [43] | 45.5 [45] | 0.99 (0.95 – 1.02) | 0.98 (0.94 – 1.02) | 0.31 |
| *Fam. Hist. of DM (%)* | 86.1 [43] | 86.7 [45] | 0.95 (0.28 – 3.21) | 0.85 (0.24 – 2.96) | 0.80 |
| *Weight (kg)* | 85.6 [44] | 78.5 [45] | 1.02 (1.00 – 1.04) | 1.03 (1.00 – 1.05) | 0.05 |
| *BMI (kg/m<sup>2</sup>)* | 30.1 [44] | 27.7 [45] | 1.05 (0.99 – 1.12) | 1.07 (1.00 – 1.15) | 0.06 |
| *A:C ratio (mg/g)* | 185.4 [44] | 40.9 [45] | 1.01 (1.00 – 1.01) | 1.00 (1.00 – 1.01) | 0.01 |
| *A:C ratio Categories:* |  |  |  |  |  |
| *Normal* | 43.18 [19] | 71.1 [32] |  |  |  |
| *Microalbuminuria* | 31.8 [14] | 24.4 [11] | 2.14 (0.81 – 5.67) | 2.21 (0.79 – 6.19) | 0.13 |
| *Macroalbuminuria* | 25.0 [11] | 4.4 [2] | 9.26 (1.85 – 46.34) | 8.35 (1.55 – 45.16) | 0.01 |
| *HbA1c % (mmol/mol)* | 9.3 (78) [44] | 8.1 (65) [45] | 1.37 (1.09 – 1.73) | 1.37 (1.08 – 1.74) | 0.01 |
| *Glucose (mg/dl)* | 151.2 [44] | 128.6 [45] | 1.01 (1.00 – 1.02) | 1.01 (1.00 – 1.02) | 0.10 |
| *Triglycerides (mg/dl)* | 98.8 [44] | 100.5 [45] | 1.00 (0.99 – 1.01) | 1.00 (0.99 – 1.01) | 0.97 |
| *Total chol (mg/dl)* | 170.3 [44] | 159.1 [45] | 1.01 (1.00 – 1.02) | 1.01 (1.00 – 1.02) | 0.28 |
| *HDL (mg/dl)* | 36.2 [44] | 34.6 [45] | 1.01 (0.98 – 1.04) | 1.00 (0.98 – 1.04) | 0.76 |
| *LDL (mg/dl)* | 112.0 [44] | 103.4 [45] | 1.01 (1.00 – 1.02) | 1.01 (1.00 – 1.02) | 0.64 |
| *VPT (volts)* | 24.5 [44] | 14.5 [45] | 1.06 (1.03 – 1.10) | 1.08 (1.03 – 1.12) | <0.01 |
| *Neuropathy (% >25V)* | 47.7 [21] | 17.8 [8] | 4.22 (1.61 – 11.10) | 4.67 (1.69 – 12.94) | <0.01 |

† Adj 1 Age, sex, diabetes duration

The demographic data in **Table 1** shows that cases (persons with current foot ulcers) and controls (persons without current foot ulcers) had the same approximate mean age (58.3 v 58.4 years respectively) and the same sex distribution (40% male). There were differences in length of diabetes duration (15.2 years in cases vs 12.8 years for controls p=0.31), age of diabetes onset (43.3 years for cases vs 45.5 years for controls; p=0.31), but these differences were not of statistical significance. Cases were heavier than controls (85.6 kg vs 78.5 kg adjusted OR 1.03; 95% CI 1.00 – 1.05 p=0.05) and had a larger mean BMI (30.1% vs 27.7% adjusted OR 1.07; 95% CI 1.00 – 1.15 p=0.06) with the differences trending toward statistical significance.

Vibration perception threshold (VPT) was markedly different between the two groups with the cases having a mean VPT of 24.5 volts vs 17.8 volts in controls (adjusted OR 1.08; 95% CI 1.03 – 1.12 p<0.01). There was also a greater proportion of cases with neuropathy (VPT >25volts) compared to controls (47.7% vs 17.8%, adjusted OR 4.67; 95% CI 1.69 – 12.94 p<0.01).

The Albumin: Creatinine ratio was different between the two groups as shown in **Table 1**. Cases had an average ratio of 185.4 mg/g while controls had a much reduced value of 40.9 mg/g, a difference that was statistically significant (OR 1.00 95% CI 1.00 – 1.01; p=0.01). The statistically significant difference was maintained when **Table 2** examined this metric with an additional regression adjustment that added the risk factors of neuropathy, body mass index and blood pressure to the regression (OR 1.01 95% CI 1.00 – 1.01; p=0.02). When subjects were stratified according to albuminuria categories, the largest proportion of cases (43%) and controls (71%) were classified as normal. In the reporting of microalbuminuria, adjusted ratios showed that cases were 3.02 times more likely than controls to be classified within the microalbuminuria (30 – 300 mg/g) range (95% CI 0.81 – 10.22 p = 0.08). In the reporting for macroalbuminuria (>300 mg/g), adjusted OR showed that cases were 7.41 times more likely than controls to have albumin: creatinine ratios in the >300mg/g range for macroalbuminuria (95% CI 1.55 – 47.49; p=0.04).

**Table 2.**
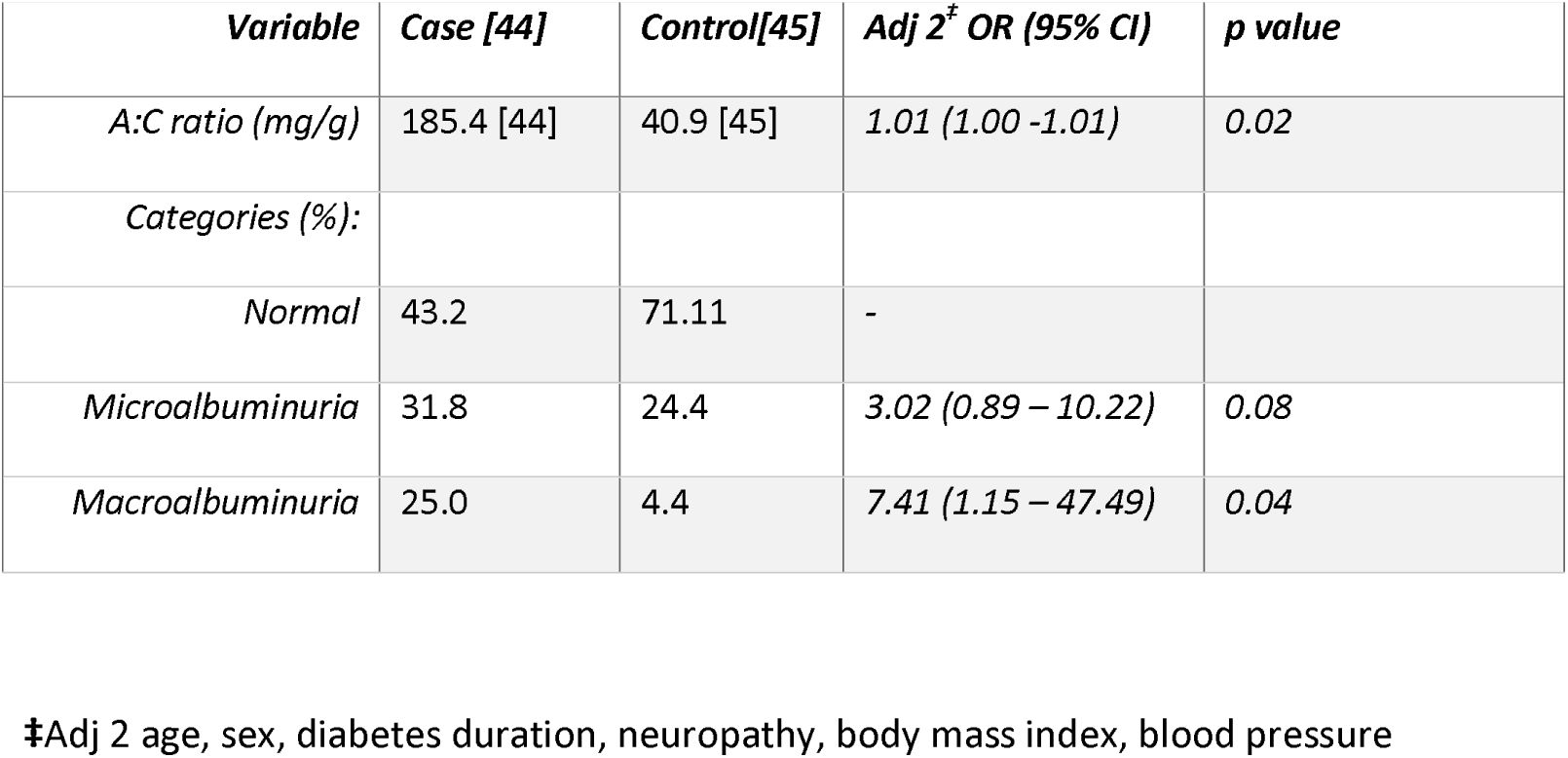
Adjusted Albumin Creatinine Odds Ratio (OR) of WHY Study Participants.

## DISCUSSION

Damage of the microvasculature of the kidneys, in particular in the glomerulus, is an underlying cause of the kidney dysfunction which manifests as microalbuminuria or macroalbuminuria or complete kidney disease. There are five stages in the progression of diabetic kidney disease as shown in **Table 3^25^** (25). The results presented in this paper show that the Albumin: Creatinine ratio was the most deranged in cases compared to controls. As shown in **Table 1** cases had a mean albumin: creatinine ratio 5 times higher than that of controls, adjusting for age, sex and duration of diabetes. When ratios were further adjusted to account for neuropathy, body mass index and blood pressure, **Table 2**, cases were 3 times more likely than controls to have microalbuminuria and 7 times more likely than controls to have macroalbuminuria. These differences were indicative of a worsened progression towards kidney malfunction in the case population compared to the control population even though none of the participants were clinically diagnosed with kidney failure or kidney disease.

**Table 3.** ^§^Classification of Diabetic Nephropathy Stages based on Urinary Albumin:Creatinine measurements. § adapted from Haneda M, Utsunomiya K, Koya D, Babazono T, Moriya T, Makino H, et al. A new Classification of Diabetic Nephropathy 2014: a report from Joint Committee on Diabetic Nephropathy. Journal of Diabetes Investigation. 2015;6(2):242-6.

| **Chronic Kidney Disease Stage** | **Definition** |
| --- | --- |
| **1 (prenephropathy)** | Normoalbuminuria (< 30 mg/g) |
| **2 (incipient nephropathy)** | Microalbuminuria (30 – 299 mg/g) |
| **3 (overt nephropathy)** | Macroalbuminuria (≥ 300 mg/g) |
| **4 (kidney failure)** | Kidney failure at any albuminuria/ proteinuria status |
| **5 (dialysis therapy)** | Any albuminuria/ proteinuria status on continued dialysis therapy |

The progression between stages of kidney disease was highlighted by Adler *et al* when it was observed that microalbuminuria develops between 5-10 years after diabetes diagnosis and that macroalbuminuria develops between 10-20 years after diabetes diagnosis^26^ (26). Research in the United Kingdom Prospective Diabetes Study (UKPDS) showed that the prevalence of microalbuminuria or worse in persons living with diabetes at diagnosis was 7.3% and rose to 24.9% after 10 years and reached 28% after 15 years across various ethnicities, confirming, through prevalence rates, the progression through the stages of diabetic kidney disease^26^ (26). Previous studies have explored an association between kidney dysfunction and diabetic foot. Some studies such as the work of Ndip et al.^27^ (27) indicated that undergoing dialysis was associated with foot ulceration in persons with stage 4 or 5 chronic kidney disease^27^ (27). Other studies have indicated that albuminuria is more present in persons with diabetic foot as well being a predictive factor of in-hospital mortality^22;23;28^ (22; 23; 28). These studies looked at persons who were at advanced stages of kidney dysfunction and kidney failure, which was in contrast to the WHY study where end stage kidney disease was not included, and dialysis was part of the exclusion criteria. By examining persons who were not in advanced kidney failure, the WHY study is therefore able to explore incipient renal compromise not possible in the aforementioned studies.

In diabetes there is a persistent low-grade inflammatory state, known as microinflammation^29^ (29). Kidney dysfunction, as evidenced in the deranged albumin creatinine ratios seen in the WHY study, may be explained by a persistent inflammatory state. Furuta demonstrated that the inflammatory state was associated with the presence of macrophages in the interstitium and glomeruli of the kidney^30^ (30). As mentioned previously, two major types of macrophages are the M1 and the M2 macrophages, with the M1 being responsible for driving the inflammatory process and the M2 having an anti-inflammatory and pro-fibrotic profile^15^ (15). In a healthy response to tissue injury, the initial M1 macrophage inflammatory phase is replaced by a M2 macrophage resolution phase, occurring 5-10 days in a model of arthropathy^31^ (31). During this switch, macrophage production of inflammatory cytokines such as IL-1β and TNF-α are suppressed whereas anti-inflammatory cytokines are upregulated, exemplified by growth factors such as TGF-β1^24; 32-33^(24; 32; 33). In animal models of acute kidney injury, the switch from M1 to M2 macrophages results in reversal of albuminuria and fewer lesions in the tubules and improved renal function^34^ (34). In chronic kidney disease however, the situation may become complicated by the prolonged presence of the M2 macrophage, since tubular interstitial fibrosis and crescent formation may result from excessive production and persistence of TGF-β secretion^35-36^ (35; 36). Landis *et al* have proposed that the worst effects of both phases may co-exist in the case of diabetic nephropathy, as a result of an incomplete transition from M1 to M2 macrophages, leading to both phenotypes being actively expressed^24^ (24). There is histological support for this “worst of both worlds” scenario, since both M1 and M2 macrophages have been detected in the glomerulus and tubulointerstitium in renal autopsy samples collected from persons with T2DM, as demonstrated by Klessens et al.^37^ (37). Klessens indicated that M1 and M2 cells coexisted in a 2:1 ratio, a scenario that may drive kidney tissue damage on the one hand by the presence of M1 macrophages and inflammatory pathways driven by AGEs, HMBGB1, reactive oxygen species and, inflammatory cytokines. On the other hand, damage is then exacerbated by basement membrane thickening and interstitial fibrosis caused by persistent release of growth factors and proteases from M2 cells. The engagement of M2 pathways may be driven by free hemoglobin and red blood cell phagocytosis pathways in the diabetic kidney microenvironment^24; 38-41^ (24; 38-41). The albumin creatinine ratios in the current study of diabetic foot, suggests that there may be a common pathophysiological pathway leading to microvascular damage underpinning impaired wound healing and diabetic nephropathy.

Neuropathy in the feet is an early step in the pathway to foot ulceration and previous studies have shown that 10-g monofilament insensitivity in the foot is associated with renal function decline^42^ (42). The comparisons between previous findings and this finding of statistically significant differences in albumin creatinine ratios between cases and controls in the WHY study are particularly interesting. They indicate that deranged albumin creatinine ratio in the Barbadian population may be an early indicator, even an early warning sentinel of a predisposition for diabetic foot ulceration which is caused in part by the dysregulation of the inflammatory process that would normally lead to wound resolution/healing.

In this paper we are highlighting the concept of a particular continuum of diabetes complications, starting with elevated plasma glucose and ending with a non-healing diabetic foot ulcer (**Figure 1)**. Based on the results of this paper, which recorded significantly deranged albumin creatinine ratio and neuropathy, as well as previous studies in diabetic foot and amputation studies in diabetes, this paper is suggesting that persons with non-healing diabetic foot ulcers are accelerated along this continuum compared to those who are diabetic but with no current foot ulcer or history of non-healing foot ulcers. The information presented in this study therefore makes a case for the use of indicators of kidney dysfunction (such as albumin creatinine ratio) as surrogate markers for the development / progression of diabetic foot in persons at increased risk of developing this complication. This is due to the fact that the microvasculature of the kidneys is more likely to manifest damage in its pathology in a manner that can be easily detected through laboratory screening, before the damage in the microvasculature of the foot can be clinically detected. In the clinical setting in Barbados, monofilaments are not used regularly and vibration perception threshold instruments are not available for use, while urine dipsticks as a proxy for kidney function are widely used. This could make screening for albuminuria more practical than screening for peripheral neuropathy. The independence of albumin: creatinine ratio in relation to other risk factors, makes it an exciting prospect in earlier identification of persons on the pathway to diabetic foot.

**Figure 1.**
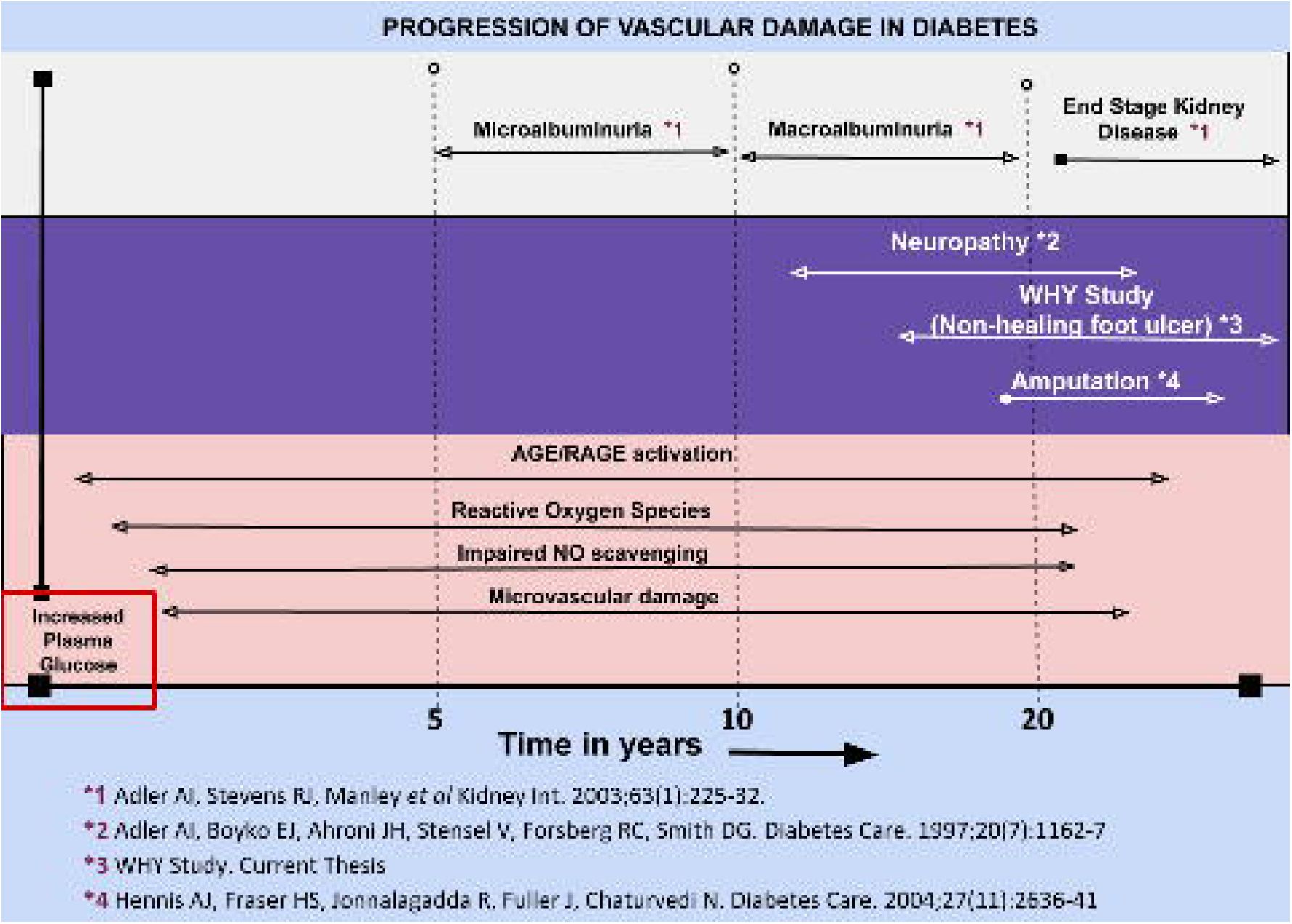
Continuum of Progression of Vascular Injury in Diabetes. This schematic illustrates the proposed timeline of progression of diabetes complications, amalgamated from various studies including the WHY study.

## Data Availability

Dr. Andre Greenidge is the guarantor of this work and, as such, had full access to all the data in the study and takes responsibility for the integrity of the data and the accuracy of the data analysis.

## ACKNOWLEDGMENTS

Funding for the WHY study was provided by the following institutions and companies: The Barbados Diabetes Foundation; Cave-Shepherd Ltd Barbados; Destiny Group of Companies, Canada; Bayer Pharmaceuticals Ltd, USA; Medicor International, UK. Core laboratory support was also received from Mr Peter Cohen and the late Mr Edmund Cohen, UK.

The study was conducted by A.G and A.S. with assistance from K.Q. All analyses were performed by A.G., I.H., and R.C.L. A.G. wrote and revised the manuscript. All authors input suggestions for analysis and interpretation and commented on the manuscript.

We thank the data team of Lourdes Soriano-Rouco and Cassandra Suttle for their work. Many thanks are also due to the Nurses in the public polyclinics and outpatient clinics of the Hospital including Sister Destang and Sister Sobers, Nurse Deborah Knight, Nurse Betty Mayers and Nurse Arlene Husbands. We also thank the members of the Barbados Diabetes Foundation, in particular Dr. Carlisle Goddard, Dr. Mike Krimholtz, Dr. Simone McConnie and the late Dr. Oscar Jordan.

The corresponding author also wishes to thank Dr Willi McFarland and Dr Sharlene Jarrett, facilitators of the **“Clear and Successful Scientific Writing Workshop”** of the University of California, San Francisco hosted by the University of the West Indies, Mona.

